# Development of a transformation model to analyze horizontal saccades using electrooculography through correlation between video-oculography and electrooculography

**DOI:** 10.64898/2026.04.14.26350920

**Authors:** Da Young Kim, Tae-Joon Kim, Yunsoo Kim, Jisu Yoo, JaeWook Jeong, Sun-Uk Lee, Jun Young Choi

## Abstract

Saccadic eye movements are established biomarkers in neuroscience and clinical neurology, where video-oculography (VOG) remains the gold standard. However, VOG’s high cost, bulky equipment, and poor portability restrict its clinical utility. Electrooculography (EOG) offers a promising alternative by detecting cornea-retinal potential changes during eye movements. To enable quantitative saccadic analysis using EOG as a VOG alternative, this study develops and validates a mathematical transformation model converting EOG data into VOG-equivalent values. A prospective observational study was conducted on 4 healthy adults without neurological or sleep disorders. Horizontal saccades were recorded simultaneously using EOG and VOG during controlled gaze shifts. EOG peak saccadic velocity was derived from voltage change rate, whereas VOG was calculated from angular displacement over time. A derivation dataset of fixed horizontal saccades (±20°) formulated the transformation model, achieving a strong correlation coefficient (r = 0.95 rightward, r = 0.93 leftward, *p* < 0.0001). Multiple filter settings were evaluated, and 0.3 Hz high-pass and 35 Hz low-pass filtering were identified as optimal. The fixed horizontal saccades derived model was applied to a validation dataset of random horizontal saccades, confirming robustness across saccades without significant differences from VOG measurements. These findings establish EOG’s feasibility for quantitative analysis of horizontal saccades and provide a validated transformation model. By systematically optimizing filtering parameters, this approach enables EOG as a cost-effective VOG alternative while maintaining high-precision measurement accuracy.

## 1. Introduction

Saccadic eye movements, characterized by rapid shifts in line of sight, serve as a crucial biomarker in various clinical and research applications [1]. Saccades consist of a hierarchy of behavior, beginning with the most fundamental eye movement—quick phases of vestibular nystagmus—through reactive saccades triggered by a sudden appearance of a novel target in the retinal periphery, to higher-level volitional behavior such as a series of saccades to scan the visual environment for new information [2]. It can be characterized by various parameters, including the waveform, velocity, duration, trajectory, and accuracy, all of which can be utilized to estimate the integrity of the neural circuit from the cerebral cortex to the brainstem [3, 4]. Saccadic analysis is employed in many fields of neuroscience, psychology, and neurological and psychiatric disease [5, 6].

Video-oculography (VOG) is a widely accepted technique for analyzing eye movements with high spatial and temporal resolution [7, 8]. However, despite its widespread adoption, its high cost and resource-intensive setup pose challenges in many clinical environments [9, 10]. In addition, the limited portability of VOG systems restricts their use in settings where patient cooperation or stable visual access cannot be ensured [11, 12]. EOG— primarily utilized in polysomnography (PSG) for sleep studies— can also reliably quantify eye movements by detecting cornea-retinal potential changes during eye movements [13, 14].

Advances in signal-processing techniques suggest that EOG can be optimized to provide reliable saccadic measurements, and its use has expanded from detection of eye opening-closing or gaze direction to more quantitative assessment of saccadic behavior [15-17]. Yet two important gaps remain. First, previous studies have not provided an extensive validation of the relationship between EOG- and VOG-derived saccadic parameters, leaving the degree to which the two modalities agree insufficiently clarified [18, 19]. Second, although HPF design critically affects EOG waveform morphology—as acausal filters can propagate later components backward in time and bias early signal segments, whereas causal filters introduce frequency-dependent attenuation that varies with cut-off frequency [20-24]—EOG-VOG comparison studies have not consistently evaluated how filter settings impact saccadic parameter estimation and EOG-to-VOG transformation stability [18, 19, 25].

To address these gaps, this study (1) performs a more extensive evaluation of the correlation between EOG and VOG peak saccadic velocities, (2) introduces a mathematically derived transformation model for converting EOG measurements into VOG-equivalent values, and (3) systematically evaluates high-pass filtering conditions to identify parameters that enable accurate EOG-based estimation of VOG equivalent peak saccadic velocities. By establishing a validated and filter-optimized mapping between EOG and VOG, this work aims to expand the clinical utility of EOG for quantitative saccadic analysis, particularly in settings where VOG is impractical or unavailable.

## 2. Methods

### 2.1. Study population

A prospective observational study involved 4 healthy individuals (two female; age range: 33-44 years) with no history of neurological or sleep disorders, all of whom provided informed consent. Ethical approval for conducting experiments on human subjects was granted by the Ajou University Hospital (Suwon, Republic of Korea, IRB no. AJOUIRB-IV-2025-253). All testing sessions were conducted at Ajou University Hospital in the Republic of Korea.

### 2.2. Data collection procedures

Eye movement was recorded using EOG from the Comet-PLUS PSG system (Grass-Technologies, Astro-Med, Inc., West Warwick, RI, USA) and VOG (SLVNG, SLMED, Seoul, Republic of Korea). Calibration of the eye position was carried out with a red dot sequentially presented from the center by 10° in vertical and horizontal directions. During the procedure, the subject was seated 1.2 m in front of the target. [26]. During 1st experiment, the subject is instructed to shift the line of gaze to a visual target that appears in horizontal direction of 20° to the left and right, repeated every 2 s. Then, the subject is instructed to shift his/her gaze to a visual target that randomly appears in time and space within 30° horizontally for 19-21 times. Monocular horizontal eye movements were recorded from the right eye, EOG was sampled at 200 Hz and VOG at 120 Hz. Both EOG and VOG signals were recorded simultaneously, resulting in two parallel data streams during the two sessions. Each recording process lasted for 40 seconds, and all tests were carried out by an instructed physician (**Fig. 1A**). Saccadic transition points in the VOG traces, produced in response to the red dot visual targets, were visually identified by a neuro-ophthalmology specialist (S.-U.L.). Time synchronization between the systems was achieved by matching the red dot onset recorded in the VOG timeline with the corresponding red dot cue in the EOG records. This procedure allowed accurate time synchronization between different sampling rates. VOG recordings were exported as *.emd* files and preprocessed using MATLAB R2023b (MathWorks Inc., Natick, MA, USA), whereas EOG recordings were exported as *.edf* files and analyzed using Python 3.11.5.

**Fig. 1.**
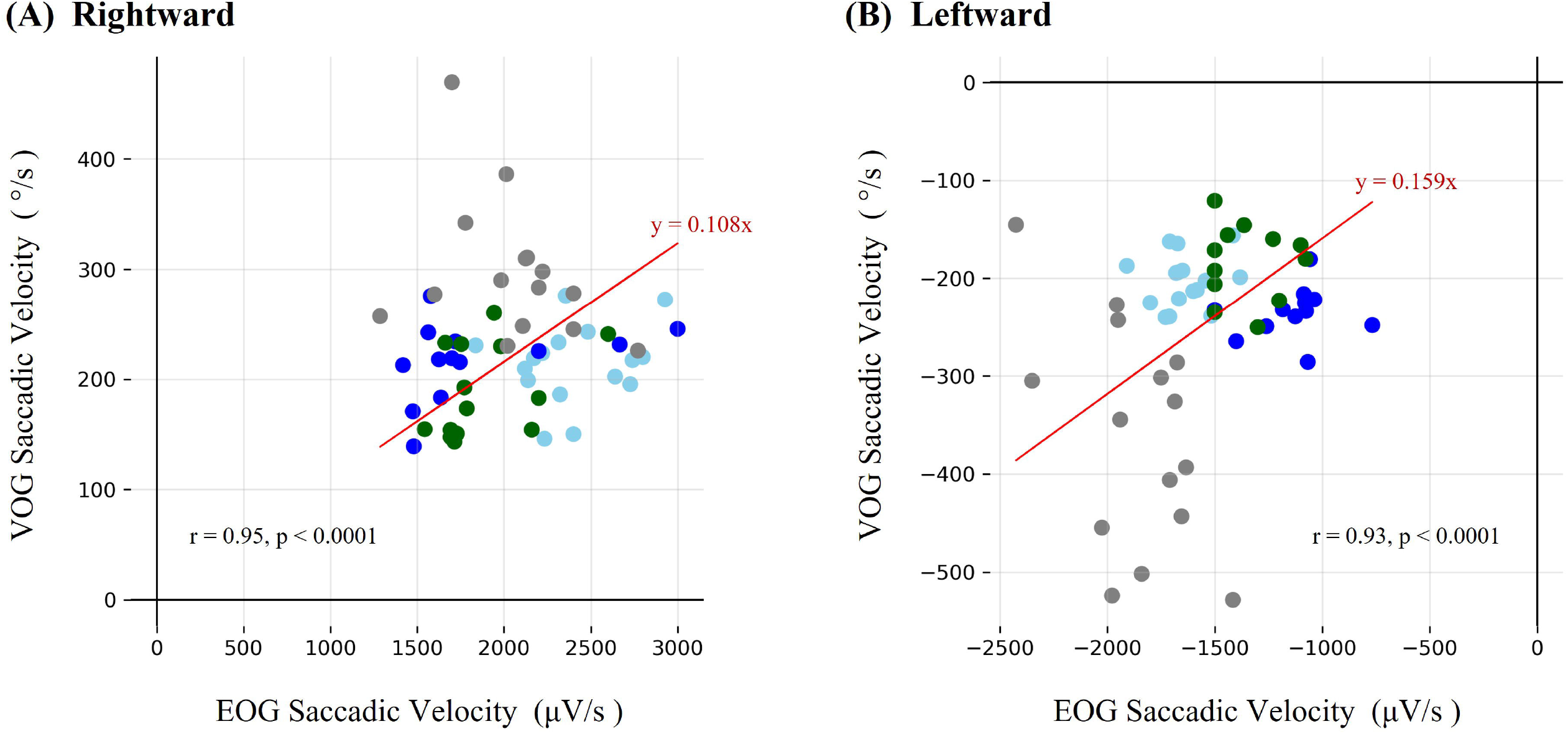
Horizontal saccadic eye movement traces and measurement schematic. (A) Horizontal saccadic eye movement traces of the right eye in a single subject (subject 1), recorded simultaneously using VOG (orange) and EOG (purple). The traces were time-synchronized to ensure accurate alignment between the two measurement modalities. (B) Schematic diagram of combined VOG and EOG measurement. The orange goggles represent the VOG device, while the purple circle indicates the location of the EOG electrode. The red dot represents the laser stimulus used to induce eye movement. d represents the initial distance between the electrode and the cornea when the eye is at rest. *r* refers to the corneal displacement caused by eye movement, and *θ* denotes the angle of the eye movement. The displacement *r* is calculated as *r*=*β*.*θ*, where *β* is a proportional constant. For *θ* ≤ 40°, the displacement *r* remains small enough to ensure *d* > *r*.

### 2.3. Signal processing and filtering

EOG signal was preprocessed using causal digital filters to examine the influence of cut-off frequency on signal morphology and EOG peak saccadic velocity estimation. To evaluate waveform distortion across different filter conditions, a synthetic EOG (−20μV to +20μV) simulating alternating rightward and leftward saccades was generated and analyzed under multiple HPF cut-off frequencies (0.1, 0.3, and 1 Hz). In addition, multiple HPF conditions were applied to the recorded EOG signals to systematically assess how each filtering cut-off frequency influenced the stability of subsequent transformation models relating to EOG and VOG peak saccadic velocities.

### 2.4. Peak saccadic velocity calculation

VOG peak saccadic velocity (*v*_*V*_): Defined as the angular velocity of the eye movement, calculated by dividing the change in gaze position (*θ*, °) by the time taken to complete <u>the movement (Δ</u>*<u>t</u>*<u>), expressed as:</u>

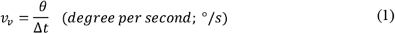

Although the correct notation is Δ*θ*, it is simplified to *θ* in subsequent equations for visual clarity and readability.

EOG peak saccadic velocity (*v*_*e*_): Defined as the rate of change in the recorded EOG signal amplitude (Δ*V*), normalized over time, expressed as:

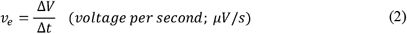

### 2.5. Statistical analysis

The uncentered Pearson correlation coefficient was used to quantify the relationship between EOG and VOG peak saccadic velocities. Unlike the standard Pearson correlation, which subtracts the mean before computing correlation, the uncentered Pearson correlation directly measures the proportional relationship between two variables [27]. This approach was chosen because the peak saccadic velocities obtained from EOG and VOG exhibited a direct proportionality passing through the origin (0, 0), making the uncentered correlation coefficient more appropriate for this analysis. A paired t-test was performed to assess the statistical significance of differences between VOG and converted EOG velocities. The P-value of < 0.05 was considered statistically significant.

## 3. Results

### 3.1. Acquisition of Peak saccadic velocity range

**Table 1** summarizes the peak saccadic velocities of the right eye measured using VOG and EOG in four subjects (number of saccades = 192), subdivided into fixed (number of saccades = 113) and random saccades (number of saccades = 79). For fixed horizontal saccades, VOG recorded a mean rightward peak saccadic velocity of 229.9 ± 60.7 °/s, with a range from 139.2 to 469.5 °/s, while leftward movements showed a higher mean velocity of −249.2 ± 96.1 °/s, ranging from −528.3 to −120.8 °/s. In contrast, EOG recorded a mean rightward velocity of 2055.3 ± 420.6 μV/s, whereas leftward movements had a lower mean velocity of −1528.6 ± 336.0μV/s. For random horizontal saccades, VOG showed a mean rightward velocity of 175.8 ± 73.3 °/s and a mean leftward velocity of −193.5 ± 106.2 °/s, while EOG recorded a mean rightward velocity of 1674.1 ± 360.4 μV/s and a mean leftward velocity of −1206.6 ± 313.4 μV/s. VOG detected leftward movements as slightly faster, whereas EOG showed that rightward movements were a little faster, suggesting that VOG and EOG may use different measurement techniques.

**Table 1.** Comparison of peak saccadic velocities of the right eye between VOG, EOG and converted EOG.

|  |  | 4 Subjects |  |  |  |
| --- | --- | --- | --- | --- | --- |
|  |  | VOG-derived | EOG-derived | Converted | P value |
|  |  | velocity | velocity | EOG-derived |  |
|  |  | (°/s) | (μV/s) | velocity |  |
|  |  |  |  | (°/s) |  |
| Horizontal Saccadic velocity |  |  |  |  |  |
| Derivation<br>(*n* = 113) | Rightward | 229.9 ± 60.7 | 2055.3 ± 420.6 | 222.0 ± 45.4 | 0.410 |
|  | Leftward | −249.2 ± 96.1 | −1528.6 ± 336.0 | −243.0 ± 53.4 | 0.638 |
| Validation<br>(*n* = 79) | Rightward | 175.8 ± 73.3 | 1674.1± 360.4 | 180.8 ± 38.9 | 0.709 |
|  | Leftward | −193.5 ± 106.2 | −1206.6 ± 313.4 | −191.9 ± 49.8 | 0.926 |

Peak saccadic velocities are shown separately for the derivation (*n* = 113) and validation (*n* = 79) datasets from four subjects. Values are presented as mean ± standard deviation (SD). Statistical comparisons between VOG-derived and converted EOG-derived velocities were conducted using paired t-tests, and the resulting p-values are reported to indicate statistical significance.

### 3.2. Development of the proposed transformation model

**Fig. 1B**. illustrates the schematic diagram of EOG measurement from an electrode placed lateral to one eye. The relationship between corneal displacement (r), eye movement angle (θ), and voltage changes was modeled as follows:

① Relationship between voltage and distance

EOG measures changes in the cornea, which varies inversely with the square of the distance (*d*) between the cornea and the recording electrode:

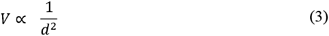

As the eye moves, the distance between the cornea and the electrode changes, resulting in a corresponding voltage change (Δ*V*). This voltage difference can be expressed as:

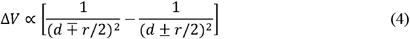

Where *d* represents the initial distance between the cornea and the electrode, and represents the displacement of the cornea due to eye movement.

② Relationship between displacement and eye movement angle

The displacement (*r*) of the cornea is proportional to the eye movement angle (θ), given by:

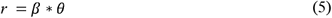

where *β* is a proportional constant.

③ Derivation of EOG and VOG velocity relationship

- VOG velocity (*v*_*V*_) was defined as the angular velocity of eye movement:

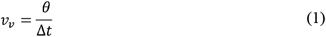
- EOG velocity (*v*_*e*_) was defined as the rate of voltage change:

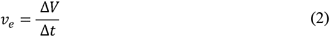

The relationship between time, angle, and voltage change is given by:

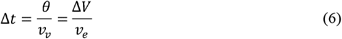

Thus, the EOG velocity can be expressed as:

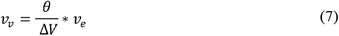

④ Substituting the displacement relationship

Incorporating the displacement equation:

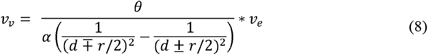

becomes,

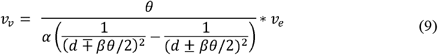

where *α* is a proportional constant.

Expanding and simplifying:

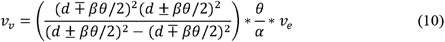

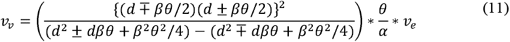

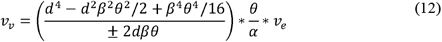

⑤ Approximation for small eye movements

Neglecting the *β*^2^*θ*^2^ term (since |*βθ*| « *d*), we obtain:

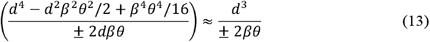

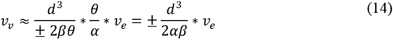

⑥ Final relationship between EOG and VOG velocities

Rearranging for *v*_*v*_, we express the relationship as:

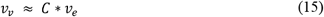

Where the proportional constant *C* differs depending on movement direction:

- Rightward saccade (+ direction):

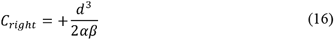

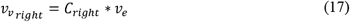
- Leftward saccade (-direction):

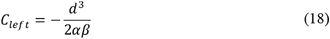

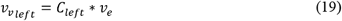

### 3.3. Filtering analysis and selection

Synthetic EOG signals were generated to evaluate HPF-induced waveform distortion and peak saccadic velocity changes across filter settings. The resulting waveform morphologies and changes in peak saccadic velocities were compared across filtering conditions (**Fig. 2**). Synthetic EOG traces demonstrated progressive waveform distortion as the HPF cut-off frequency increased. Consistent with the experimental recordings, both synthetic and real EOGs showed a gradual reduction in peak saccadic velocity at higher HPF cut-offs (**Table S1**).

**Fig. 2.**
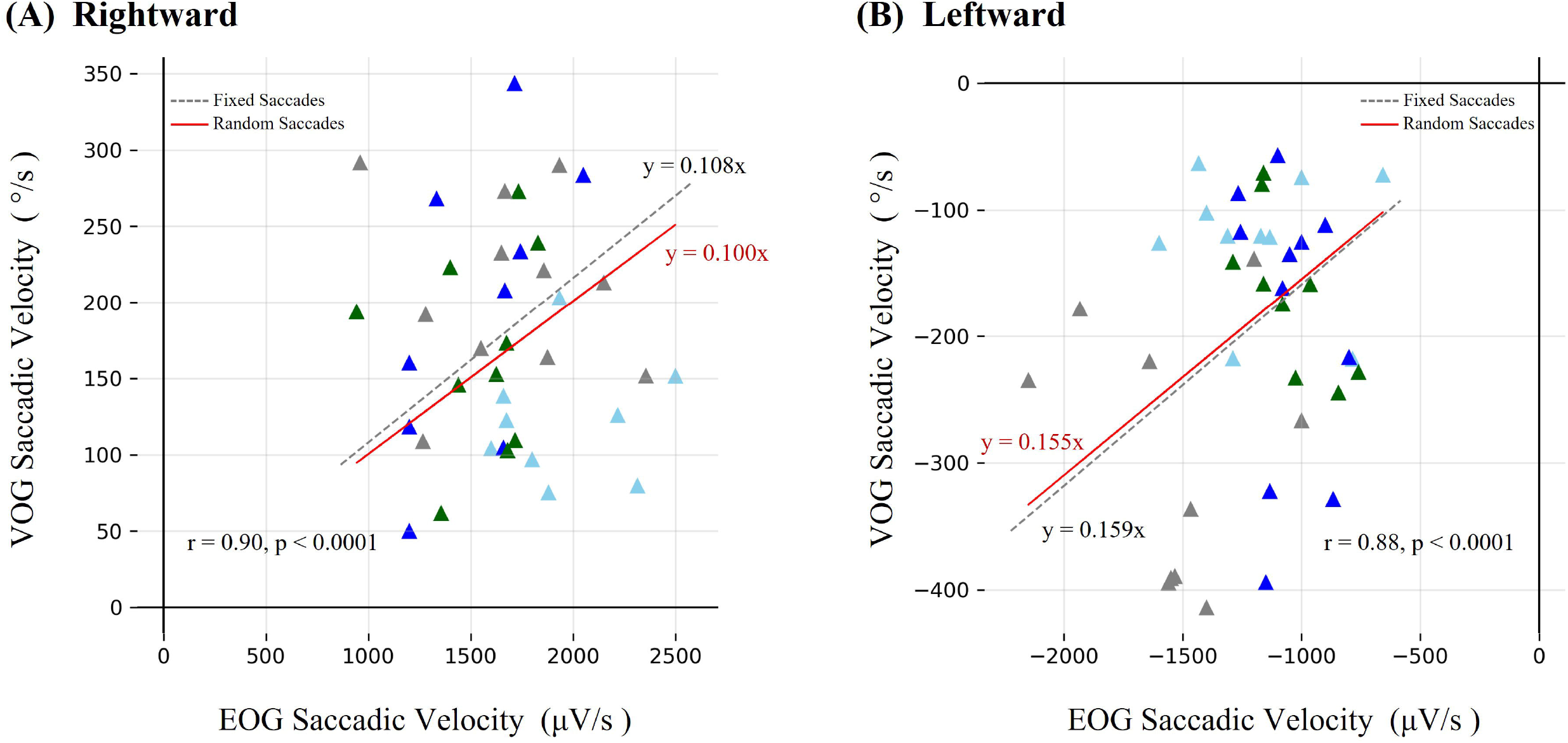
Comparison of saccadic velocities under High-pass filter (HPF) cut-offs on signal waveform in a synthetic EOG. Rapid change points 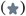 denote saccade onset, followed by right-gaze 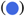 or left-gaze 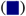 fixation. Saccadic velocity represents the voltage change from onset to each fixation target. All synthetic EOG waveforms were processed with a fixed low-pass filter (35 Hz). Gaussian noise and baseline drift were added to emulate real-world EOG characteristics.

Based on these preliminary observations, multiple HPF conditions were applied to the recorded EOG, and transformation models were estimated from derivation dataset (fixed horizontal saccades, ± 20°) under each filtering condition (**Fig. S1**). The resulting direction-specific regression constants were then tested in the validation dataset of random horizontal saccades. For each filter setting, the agreement between converted EOG and VOG velocities and consistency of regression slopes across datasets were evaluated (**Fig. S2**). Supplementary analysis across different HPF cut-off frequencies demonstrated no significant differences between VOG-derived and converted EOG-derived velocities (**Table S2**,**3**), indicating that transformation model remained robust across the tested HPF settings.

In parallel, several low-pass filter (LPF) cut-off frequencies were examined to assess their impact on peak saccadic velocity estimates. Exploratory analysis showed that an LPF of 35 Hz yielded the most consistent agreement between derivation and validation datasets, supporting its use as a practical LPF setting for the proposed transformation model (**Fig. S3**). Taken together, the HPF 0.3 Hz and LPF 35 Hz configuration provided the most stable and directionally consistent performance and was therefore selected as the optimal filtering configuration for subsequent analysis.

### 3.4. Correlation analysis

The mathematical model was derived using experimental data. **Fig. 3**. presents scatter plots illustrating the correlation between EOG peak saccadic velocity (*µV*/*s*) and VOG peak saccadic velocity (*degree*/*s*) for both rightward and leftward saccades of the right eye. The following regression equations were derived from the fixed horizontal saccades dataset under the optimal filtering condition.

**Fig. 3.**
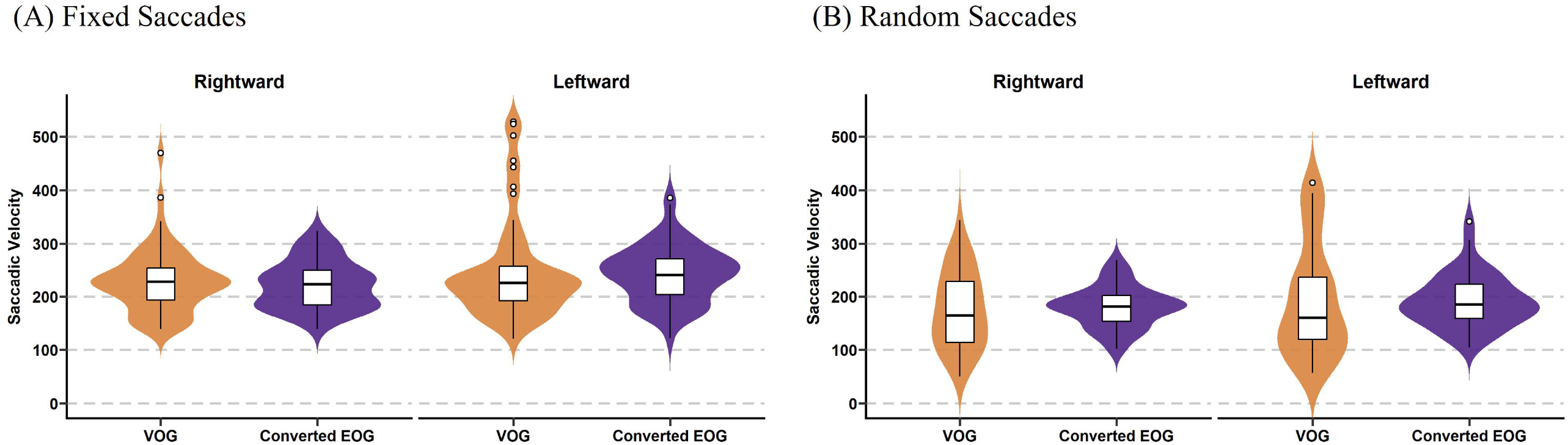
Correlation of EOG and VOG saccadic velocity of the right eye in derivation dataset (fixed saccades). (A) Rightward; (B) leftward. Data points for Subject 1 (sky blue), subject 2 (blue), subject 3 (green), and subject 4 (gray) are shown. The “●” symbol represents a fixed 40° position. The red line represents the linear relationship between EOG and VOG saccadic velocity, fitted without an intercept (y = ax), reflecting a direct proportionality between the two variables. The uncentered Pearson correlation coefficient (r) was calculated to quantify the strength and direction of the linear relationship, reflecting the proportional relationship between EOG and VOG saccadic velocity without adjusting for the mean of each variable. The associated p-value (p) is provided to assess the statistical significance of the observed correlation.

- Rightward saccade (+direction): The regression equation is

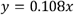

with a correlation coefficient is *r*=0.95, with a highly significant p-value (*p* <0.0001).
- Leftward saccade (−direction): The regression equation is

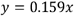

with *r*=0.93 and *p*<0.0001, similarly demonstrating a strong linear relationship.

These constants *C* from fixed horizontal saccades (Fig. 3, derivation dataset) were directly applied to the random horizontal saccades (validation dataset) without modification. Fig. 4 demonstrates the correlation analysis results, where gray dashed lines represent the derivation-derived constants *C*, and solid red lines show newly fitted slopes from validation data. Consistent linear relationships across both datasets confirm the generalizability of the EOG-to-VOG transformation model.

**Fig. 4.**
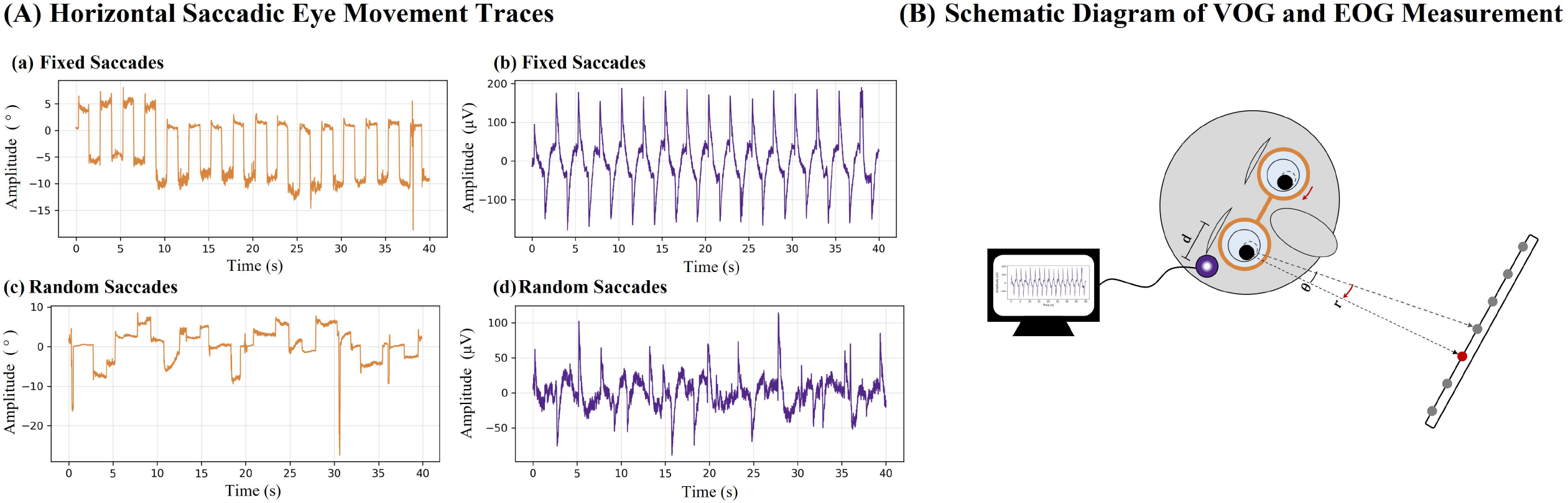
Correlation of EOG and VOG saccadic velocity of the right eye in the validation dataset (random saccades). (A) Rightward; (B) leftward. Data points for Subject 1 (sky blue), subject 2 (blue), subject 3 (green), and subject 4 (gray) are shown. The “▴” symbol represents random saccades. The red line represents the linear relationship between EOG and VOG saccadic velocity in validation dataset, fitted without an intercept (y = ax), whereas gray dashed line represents the corresponding regression line derived from the derivation dataset shown in Fig. 3. As in Fig. 3, the uncentered Pearson correlation coefficient (r) and p-value (p) are shown in each panel to indicate the strength and statistical significance of the observed linear relationship.

Under physiologically appropriate filtering conditions (0.3 Hz high-pass, 35 Hz low-pass), the proposed transformation model produced regression constants that remained stable between derivation (Fig. 3) and validation (Fig. 4) datasets, demonstrating the reliability of EOG-based measurements for quantifying saccadic eye movements.

### 3.5. Comparison of VOG velocity and converted EOG velocity

After applying the transformation model, no statistically significant differences were observed between the converted EOG velocities and the VOG velocities (**Table 1**). This result supports the validity of the proposed transformation model and confirms that EOG-based measurements can be reliably converted into VOG-equivalent values in both derivation (fixed horizontal saccades) and validation (random horizontal saccades). The comparison is further illustrated in **Fig. 5**., where the distributions of VOG and converted EOG velocities are closely aligned for representative filter setting (HPF 0.3 Hz, LPF 35 Hz), and **Fig. S4**, which shows similar alignment across multiple HPF conditions with LPF fixed at 35 Hz. The statistical analysis and visualization collectively indicate that the proposed transformation model effectively aligns EOG-derived velocities with VOG measurements.

**Fig. 5.**
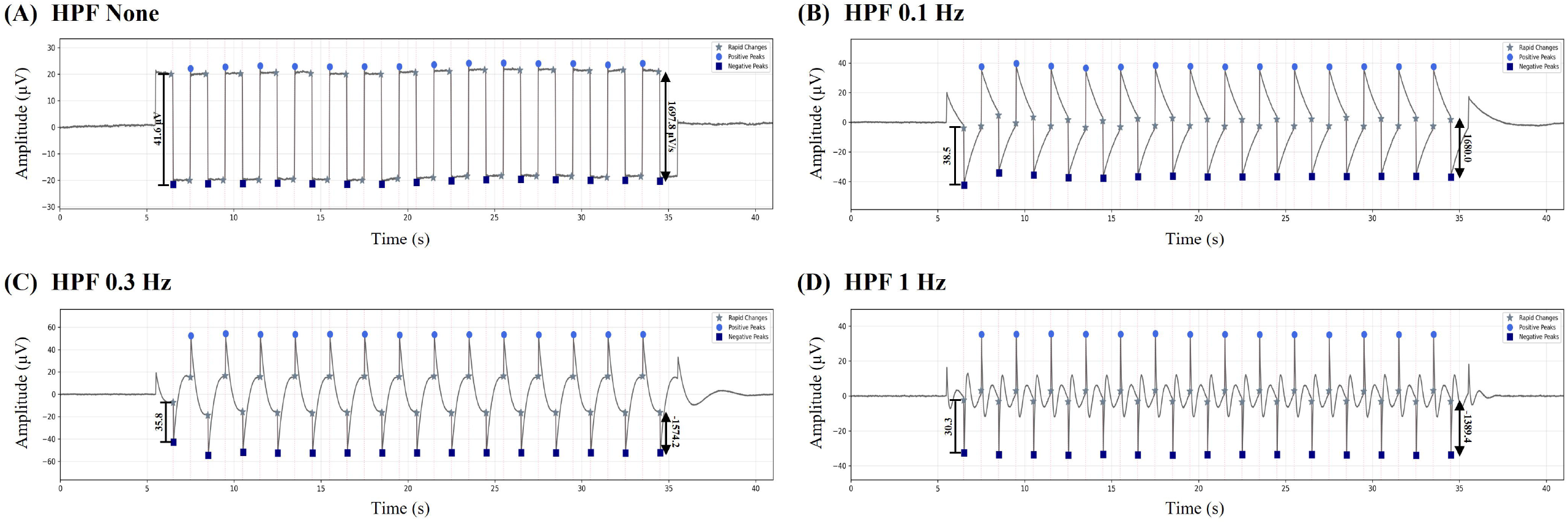
Violin plots comparing peak saccadic velocities of the right eye between VOG and converted EOG (using transformation model) for (A) fixed saccades and (B) random saccades. Each violin plot shows the full distribution of velocities; central white box = interquartile range (IQR), horizontal line = median, and outer shapes = probability density.

Furthermore, supplementary analysis across multiple HPF cut-off frequencies (see **Table S2**,**3**) consistently showed no significant differences between VOG and converted EOG velocities, reinforcing the robustness and generalizability of the proposed transformation model under various filtering conditions.

## 4. Discussion

### 4.1. Significance of the study

This study provides a quantitative foundation for utilizing EOG in saccadic eye movement assessments. By developing a robust mathematical transformation model, it enables the conversion of EOG-derived peak saccadic velocities into VOG-equivalent values, enhancing the clinical utility of EOG. In clinical settings, VOG is typically performed while sitting and requires substantial cooperation from the subject. Consequently, systematic measurement of eye movement is limited in patients who are bedridden or who have altered mentality. This study highlights the potential of the EOG as a more accessible and cost-effective solution that allows multimodal signal processing captured during EEG or PSG. These findings pave the way for broader applications of EOG in both clinical diagnostics and research.

### 4.2. Peak saccadic velocity comparison

Prior studies have reported that, at a 30 deg amplitude, average horizontal peak saccadic velocities exceed 200 deg/s in normal subjects [28]. Similarly, our study produced comparable results, with peak saccadic velocities consistently falling within these reference ranges.

A discrepancy was found between EOG and VOG measurements of peak saccadic velocities in the right eye. VOG recorded faster leftward movements, while EOG showed the opposite trend. This asymmetry in EOG may be due to electrode placement on the right side. As the eye moves rightward, the cornea approaches the electrode, producing a stronger voltage shift and higher velocity readings. Conversely, leftward movements increase the distance from the electrode, resulting in lower recorded velocities. This placement effect likely contributed to the difference between EOG and VOG measurements.

### 4.3. Correlation and validation of the EOG to VOG transformation model

The study revealed a strong correlation between EOG and VOG peak saccadic velocities, with correlation coefficients of *r*= 0.95 for rightward saccades and *r*= 0.93 for leftward saccades in the derivation set. These high correlation values underscore the reliability of EOG as a viable alternative to VOG when combined with the proposed transformation model.

The proposed transformation model effectively converted EOG-derived velocities into VOG-equivalent values. The mathematical model was derived based on the relationship between corneal displacement, eye movement angle, and voltage changes. The transformation model incorporated proportional constants that adjusted for directional differences during saccades.

Statistical comparisons between VOG and converted EOG velocities showed no significant differences, demonstrating that the model estimates VOG-equivalent velocities from EOG data. By enabling the precise conversion of EOG-derived velocities, this transformation model enhances the clinical and research utility of EOG-based saccadic assessments.

### 4.4. Directional asymmetry in transformation constants

Although the proposed transformation model predicts identical regression constants for rightward and leftward saccades, the constants derived from experimental data differed (*C*_*left*_ > *C*_*right*_). This asymmetry likely arises from three systematic factors. First, slight differences in electrode placement and dipole-to-electrode geometry alter the effective sensitivity to corneal approach versus withdrawal, resulting in unequal voltage-distance scaling [19]. Second, recording system characteristics include high-pass filtering, amplifier phase response, and electrode skin impedance introduce frequency-dependent attenuation that affects the positive and negative EOG transients differently, producing direction-specific velocity scaling. Third, the simplified dipole model assumes symmetric 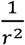 decay and purely rotational motion, whereas real eye movements include translational components and anisotropic tissue conduction, leading to deviations from ideal symmetry [18]. Fourth, study-specific factors may have also contributed to the observed asymmetry: The physical frame of the VOG goggles may have influenced the EOG signal, the EOG electrode may not have been positioned perfectly horizontally but slightly obliquely, and the recorded EOG signal may have included contributions not only from the right cornea but also from the contralateral cornea. These factors collectively explain why *C*_*right*_ and *C*_*left*_ diverged despite theoretical expectations, indicating that direction-specific calibration may further improve EOG-to-VOG transformation accuracy.

### 4.5. Clinical and research implications

The strong correlation observed between EOG and VOG peak saccadic velocities underscore the feasibility of utilizing EOG for saccadic assessments. VOG has been widely recognized as the gold standard for eye movement analysis due to its ability to precisely measure angular displacement and velocity [7]. However notable limitation of VOG is its reliance on video-based tracking, which inherently requires a clear and continuous visual access to the pupils in order to accurately track gaze direction. Moreover, VOG systems lack portability, making their use challenging in intensive care units (ICUs), emergency settings, or for patients who are bedridden, comatose, or otherwise unable to cooperate. This restriction renders VOG ineffective in conditions where eye visibility is compromised, such as when subjects have ptosis, severe eyelid abnormalities, or are in a sleep state [11]. In contrast, EOG operates independently of eye visibility, making it particularly advantageous for applications requiring continuous eye movement monitoring [29]. Furthermore, the cost and complexity of VOG systems pose significant barriers to widespread adoption, particularly in resource-limited environments. EOG, being more cost-effective and easier to implement, presents a practical alternative for conducting in diverse clinical settings. This suggests that EOG, when calibrated using the proposed transformation model, may serve as a viable tool for assessing saccadic eye movements in non-cooperative or critically ill patients.

### 4.6. Limitations and future directions

Despite its strengths, this study has certain limitations that warrant further investigation. First, the study was conducted on only four healthy participants. These findings warrant further validation in larger populations to determine whether the diagnostic yield of the transformational model remains consistent across individuals. Additionally, future research can be adopted in patients with neurological disorders affecting the saccadic system to validate the model’s generalizability. Second, this study focused on controlled horizontal saccadic eye movements. Future research should examine whether the transformation model remains valid under more dynamic and complex visual tracking conditions, such as free-viewing tasks. Third, horizontal eye movements were recorded monocularly from the right eye. Although this may have appeared to be a methodological limitation, the bilateral symmetry of horizontal ocular dipole orientation and saccadic mechanics suggests that EOG signals recorded from left eye would exhibit a mirror-symmetric pattern. Therefore, analyzing only EOG from the right eye was considered sufficient for evaluating the transformation model, although future studies should confirm this symmetry explicitly.

## 5. Conclusion

This study establishes a strong correlation between EOG and VOG peak saccadic velocities and introduces a mathematical transformation model that enables the conversion of EOG-derived data into VOG-equivalent values. As the first study to quantitatively validate this relationship, these findings reinforce the feasibility of using EOG as a cost-effective and accessible alternative to VOG, particularly in settings where VOG is impractical due to cost, equipment requirements, or patient constraints. By expanding the applicability of EOG-based eye movement analysis, this research provides a foundation for its integration into broader clinical and research applications.

## Credit authorship contribution statement

**Da Young Kim:** Writing – review & editing, Visualization, Software, Formal analysis. **Tae-Joon Kim:** Conceptualization, Resources, Methodology, Writing – original draft, Writing – review & editing, Funding acquisition. **Yunsoo Kim:** Investigation, Writing – original draft. **Jisu Yoo:** Investigation, Data curation. **JaeWook Jeong:** Data curation, Software. **Sun-Uk Lee:** Validation, Supervision, Data curation, Writing – original draft, Writing – review & editing. **Jun Young Choi:** Resources, Conceptualization, Supervision, Methodology, Writing – original draft, Writing – review & editing, Funding acquisition.

## Data availability

Data will not be made publicly available to preserve the privacy of the research participants.

