## Supplementary Tables and Figures for "Development of a transformation model to analyze horizontal saccades using electrooculography through correlation between video-oculography and electrooculography"

**Correlation of Peak Saccadic Velocities between Electrooculography and Video-oculography**

**Supplementary Figure**

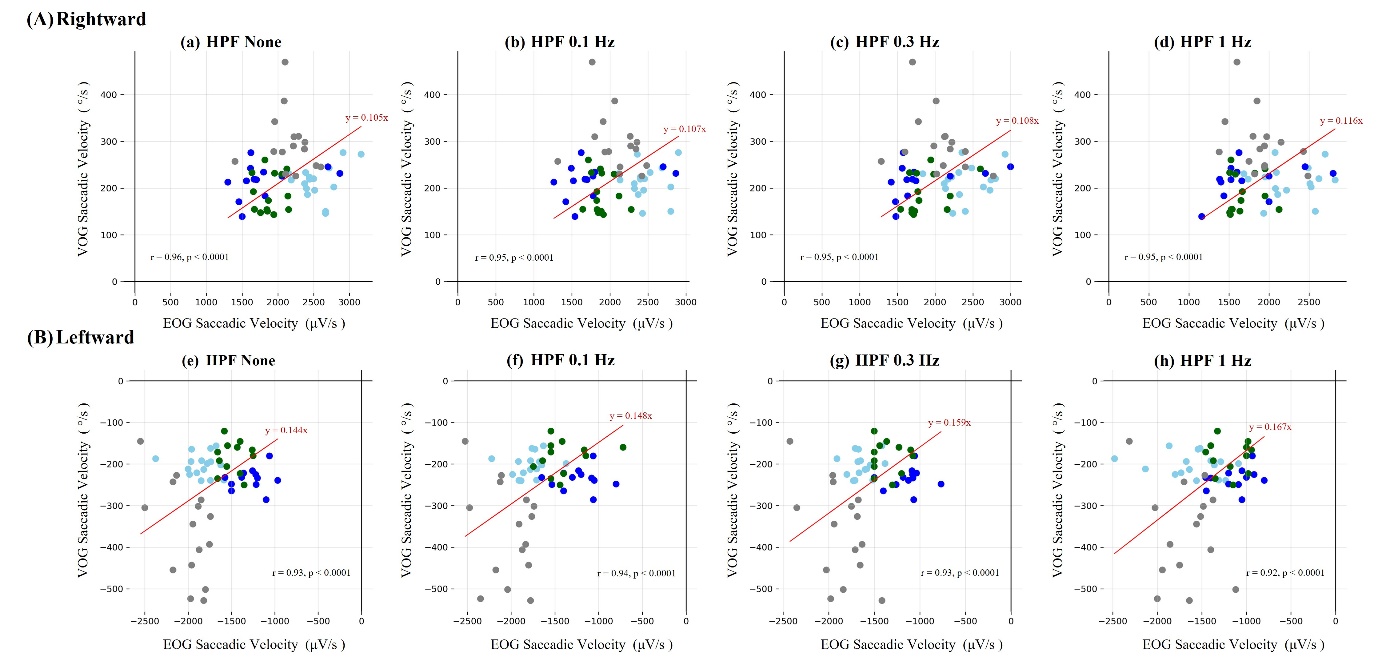

**Fig. S1.** Correlation of EOG and VOG peak saccadic velocities of the right eye according to HPF frequency in the derivation dataset. (A) Rightward; (B) leftward. Data points for Subject 1 (sky blue), subject 2 (blue), subject 3 (green), and subject 4 (gray) are shown. The “●” symbol represents a fixed 40° position. The red line in each panel represents the linear fit without intercept (y = ax). Uncentered Pearson correlation coefficient (r) and their associated p-values (p) quantify the strength and statistical significance of the proportional relationship between EOG and VOG peak saccadic velocities.

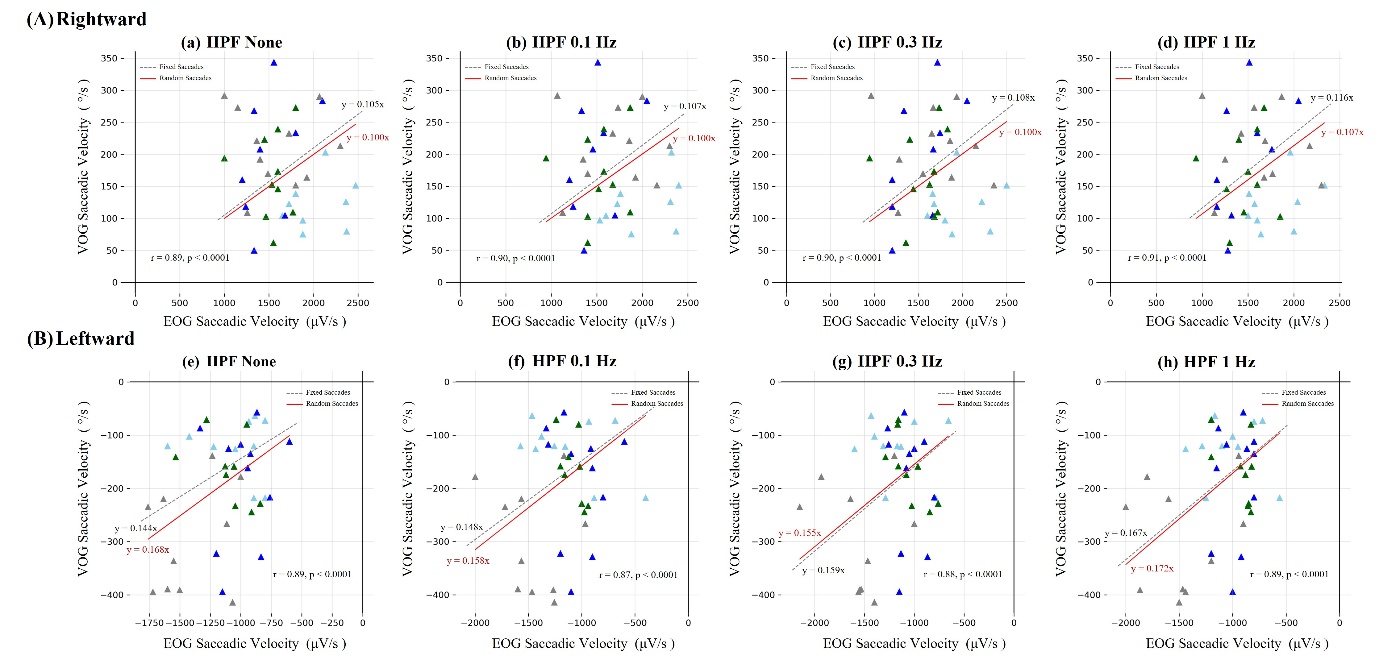

**Fig. S2.** Correlation of EOG and VOG peak saccadic velocities of the right eye in random saccades. (A) Rightward; (B) leftward. The “▲” symbol represents random saccades. The solid red line shows the regression line for random saccades (y = ax), while the gray dashed line represents the fixed saccade regression from Fig. S1 for comparison. The uncentered Pearson correlation coefficients (r) and p-values (p) indicate the strength and significance of the proportional relationship between EOG and VOG saccadic velocities.

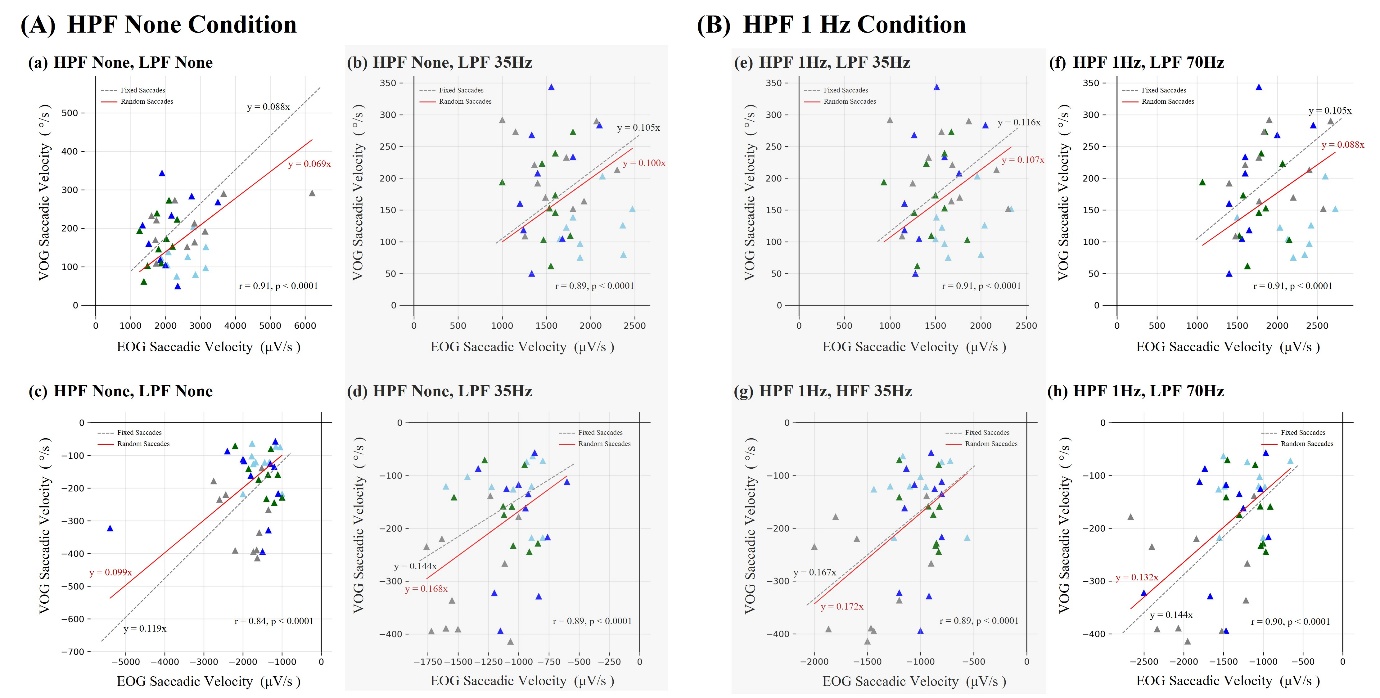

**Fig. S3.** Correlation between EOG and VOG saccadic velocities of the right eye under different LPF cut-off frequencies. (A) HPF None condition comparing LPF None and 35 Hz (panels a-d), and (B) HPF 1 Hz condition comparing LPF 35 Hz and 70 Hz (panels e-h). The top two rows represent rightward saccades, and the bottom two rows represent leftward saccades. Gray dashed lines indicate regressions for fixed saccades, and red solid lines indicate regressions for random saccades. Closer agreement between the gray dashed (derivation) and red solid (validation) regression lines indicates better consistency of the transformation constant *C* between the derivation and validation datasets.

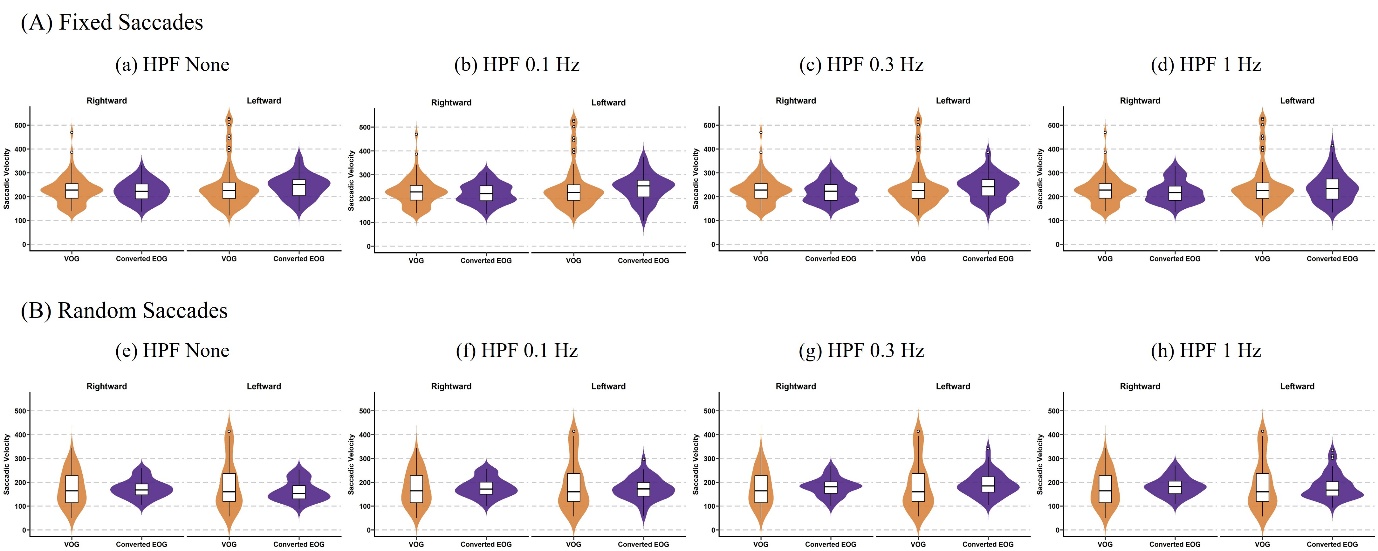

**Fig. S4.** Violin plots comparing peak saccadic velocities of the right eye between VOG and converted EOG (using the transformation model) under different HPF conditions, with LPF fixed at 35 Hz. For both (A) fixed and (B) random saccades, each violin shows the distribution of velocities; the central white box represents the interquartile range (IQR), and the horizontal line inside the box indicates the median velocity.

**Supplementary Table**

**Table S1**

Comparison of saccadic velocities of the right eye between synthetic and real-world EOG signals across different HPF settings

|  | | Synthetic EOG | |  | Real-world EOG | |
| --- | --- | --- | --- | --- | --- | --- |
|  | | Horizontal Saccadic velocity (μV/s) | Relative to HPF None (%) |  | Horizontal Saccadic velocity (μV/s) | Relative to HPF None (%) |
| HPF None | |  |  |  |  |  |
|  | Rightward | 2061.7 ± 13.8 | 100.0 |  | 2127.2 ± 419.4 | 100.0 |
|  | Leftward | $-$1693.2± 7.7 | 100.0 |  | −1687.2 ± 348.7 | 100.0 |
| HPF 0.1 Hz | |  |  |  |  |  |
|  | Rightward | 2043.9 ± 13.5 | 99.1 |  | 2073.4 ± 401.2 | 97.5 |
|  | Leftward | $-$1675.9 ± 8.9 | 99.0 |  | −1645.6 ± 389.0 | 97.5 |
| HPF 0.3 Hz | |  |  |  |  |  |
|  | Rightward | 1934.2 ± 14.4 | 93.8 |  | 2055.3 ± 420.6 | 96.6 |
|  | Leftward | $-$1575.2 ± 8.8 | 93.0 |  | −1528.6 ± 336.0 | 90.6 |
| HPF 1 Hz | |  |  |  |  |  |
|  | Rightward | 1739.9 ± 13.2 | 84.4 |  | 1907.0 ± 404.7 | 89.6 |
|  | Leftward | $-$1390.2 ± 9.6 | 82.1 |  | −1426.6 ± 369.9 | 84.6 |

Values are presented as mean ± standard deviation (SD). Absolute Saccadic velocity progressively decreased as the high-pass filter (HPF) cut-off frequency increased from none to 1 Hz, showing a similar trend in both synthetic and real-world EOG signals.

**Table S2**

|  |  | 4 Subjects (*n* = 113) | | |  |
| --- | --- | --- | --- | --- | --- |
|  |  | VOG-derived velocity  (°/s) | EOG-derived velocity  (μV/s) | Converted EOG-derived velocity  (°/s) | P value |
| Horizontal  Saccadic velocity |  |  |  |  |  |
| HPF None | Rightward | 229.9 ± 60.7 | 2127.2 ± 419.4 | 223.4 ± 44.0 | 0.475 |
|  | Leftward | $-$249.2 ± 96.1 | −1687.2 ± 348.7 | $-$243.0 ± 50.2 | 0.637 |
| HPF 0.1 Hz | Rightward | 229.9 ± 60.7 | 2073.4 ± 401.2 | 221.9 ± 42.9 | 0.406 |
|  | Leftward | $-$249.2 ± 96.1 | −1645.6 ± 389.0 | $-$243.6 ± 57.6 | 0.655 |
| HPF 0.3 Hz | Rightward | 229.9 ± 60.7 | 2055.3 ± 420.6 | 222.0 ± 45.4 | 0.410 |
|  | Leftward | $-$249.2 ± 96.1 | −1528.6 ± 336.0 | $-$243.0 ± 53.4 | 0.638 |
| HPF 1 Hz | Rightward | 229.9 ± 60.7 | 1907.0 ± 404.7 | 221.2 ± 46.9 | 0.384 |
|  | Leftward | $-$249.2 ± 96.1 | −1426.6 ± 369.9 | $-$238.3 ± 61.8 | 0.431 |

Comparison of saccadic velocities of the right eye between VOG, EOG and Converted EOG in the derivation dataset under different HPF conditions.

Values are presented as mean ± standard deviation (SD). Statistical comparisons between VOG-derived velocity and converted EOG-derived velocity were conducted using paired t-tests, and the resulting p-values are reported to indicate statistical significance. These results were obtained from a total of 113 velocity measurements from four participants in the derivation dataset.

**Table S3**

Comparison of saccadic velocities of the right eye between VOG, EOG and Converted EOG in the validation dataset under different HPF conditions.

|  |  | 4 Subjects (*n* = 79) | | |  |
| --- | --- | --- | --- | --- | --- |
|  |  | VOG-derived velocity  (°/s) | EOG-derived velocity  (μV/s) | Converted EOG-derived velocity  (°/s) | P value |
| Horizontal  Saccadic velocity |  |  |  |  |  |
| HPF None | Rightward | 175.8 ± 73.3 | 1660.6 ± 369.9 | 174.4 ± 38.8 | 0.919 |
|  | Leftward | $-$193.5 ± 106.2 | $-$1130.9 ± 295.2 | $-$162.8 ± 42.5 | 0.061 |
| HPF 0.1 Hz | Rightward | 175.8 ± 73.3 | 1670.4 ± 374.4 | 178.7 ± 40.1 | 0.829 |
|  | Leftward | $-$193.5 ± 106.2 | $-$1169.0 ± 315.3 | $-$173.0 ± 46.7 | 0.247 |
| HPF 0.3 Hz | Rightward | 175.8 ± 73.3 | 1674.1± 360.4 | 180.8 ± 38.9 | 0.709 |
|  | Leftward | $-$193.5 ± 106.2 | $-$1206.6 ± 313.4 | $-$191.9 ± 49.8 | 0.926 |
| HPF 1 Hz | Rightward | 175.8 ± 73.3 | 1581.7 ± 339.8 | 183.5 ± 39.4 | 0.560 |
|  | Leftward | $-$193.5 ± 106.2 | $-$1100.2 ± 329.1 | $-$183.7 ± 55.0 | 0.539 |

Values are presented as mean ± standard deviation (SD). Statistical comparisons between VOG-derived velocity and converted EOG-derived velocity were conducted using paired t-tests, and the resulting p-values are reported to indicate statistical significance. These results were obtained from a total of 79 velocity measurements from four participants in the derivation dataset.
